# Integrative Transcriptomic Analysis Reveals Molecular Signatures and Candidate Therapeutic Targets for Primary Ciliary Dyskinesia

**DOI:** 10.64898/2026.01.12.26343910

**Authors:** Jitender, Md Wamique Hossain, Shilpa Mohanty, Suneel Kateriya

## Abstract

Primary ciliary dyskinesia (PCD) is a rare genetic disorder arising from motile cilia dysfunction, causing chronic respiratory infections, bronchiectasis, and progressive lung damage. We analysed the GSE25186 microarray dataset (6 PCD vs. 9 controls, Illumina HumanHT-12 V3.0, GPL6947) using an empirical-Bayes moderated t-test with sex-covariate correction. After Benjamini-Hochberg adjustment, 176 genes were differentially expressed (nominal p < 0.01, |log2FC| > 1; no gene reached FDR < 0.05 after multiple-testing correction), with a broader set of 1,014 genes at nominal p < 0.05. After sex correction, biologically coherent PCD-relevant genes (MED13L, DDX58, HLA-C, CLN8, TP53BP2, SLC18A1) emerged at the top of the ranked list. MED13L is a mediator complex subunit that is involved in FOXJ1-driven ciliogenesis. MED13L emerged as the top network hub gene. The WGCNA was unstable at n = 15 (standard requirement >= 20 samples). Therefore, module-trait correlations did not reach significance after permutation testing (all permutation p > 0.05). A fully nested leave-one-out cross-validation pipeline yielded AUC = 0.750 (95% CI 0.43-1.00, permutation p = 0.062). LINCS L1000 connectivity analysis identified curcumin and resveratrol as top repurposing candidates; NAC was not significant (p = 0.37). Molecular docking against MED13L (AlphaFold, CDK8-interaction domain) ranked dexamethasone (−6.2 kcal/mol) and resveratrol (−5.9 kcal/mol) above NAC (−3.7 kcal/mol). The results demonstrate the critical importance of sex-covariate correction, nested cross-validation, and cross-cohort replication in small-cohort rare-disease transcriptomics.

## 1. Introduction

Primary ciliary dyskinesia (PCD) belongs to a group of heterogeneous diseases that are characterized by motile cilia dysfunction. Being a rare condition, it is roughly estimated to exist 1 in every 15,000 or 20,000 live births [1, 2]. However, this assessment can be underrepresented due to limitations in diagnostics and high variability in genetics [3]. The clinical manifestations of PCD include respiratory distress, rhinosinusitis, chronic productive cough, recurring pneumonia, and progressive bronchiectasis. These symptoms may also lead to complete respiratory collapse, often leading to lung transplantation in severe cases [4, 5]. However, the symptoms of PCD are not limited to respiratory distress. In 50 % of the cases, it often leads to laterality defects, including complete situs inversus totalis, and heterotaxy syndromes. These broad-spectrum phenotypes highlight the crucial role of embryonic nodal cilia in determining the left-right symmetry [6]. It also manifests as a reproductive organ dysfunction, as it causes male infertility in almost 100 % of the PCD patients. Its wide-ranging effects also include conductive hearing loss due to persisting otitis media with effusion and retinitis pigmentosa. Further, in rare cases, it exhibits hydrocephaly [7].

Studies involving PCD have identified over 50 genes that have been implicated in the development of PCD. However, it represents only 70 % of the cases, indicating that several more causative genes need to be discovered [8]. CCDC39 and CCDC40 constitute the inner dynein arm (IDA) components that result in 12-18 % of the IDA-related defects [9, 10]. In occasional cases (approximately 2-3 %), PCD manifests due to radial spoke proteins (RSPH1, RSPH4A, RSPH9) dysfunction that leads to intermittent or impaired cilia dysfunction [11]. The rarest forms of PCD exist due to mutations in the central apparatus components, including HYDIN and SPEF2, that often lead to ciliary beat pattern dysfunction. An abnormal beat pattern depicts that structural analysis alone is not sufficient to diagnose PCD [12]. The improper assembly of inner and outer dynein arms can occur in 15-20 % of cases, which often arises due to defects in cytoplasmic assembly factors such as DNAAF1-5, LRRC6, ZMYND10, and C21orf59 [13, 14]. Recent studies have demonstrated that reduced ciliary dysfunction can also occur due to reduced generation of multiple motile cilia (RGMC), where multiple transcriptional factors such as FOXJ1, MCIDAS, and CCNO are implicated [15].

The nasal nitric oxide measurement test has failed in diagnosing PCD due to the lack of specificity. Further, it may show normal values in a few genotypes, particularly with RSPH1 mutations [4]. High-speed video microscopy is incapable of reliably detecting abnormal beating or static cilia with residual motion and requires specialized expertise [16, 17]. Transmission Electron Microscopy (TEM) is capable of identifying only 70 % of the PCD cases, implying the lack of normal ultrastructural variants. This necessitates multiple biopsies for efficient diagnosis, which is time-consuming and expensive [18].

Since PCD belongs to the group of rare diseases, it is limited by the availability of a small sample size and limited resources. Hence, integrative computational approaches become more prominent for such cases for extracting maximum biological insights from the accessible data [19, 20]. Several transcriptomic studies have been carried out for PCD; however, they lack depth and scope. Geremek and co-workers carried out expression profiling studies on respiratory epithelial cells isolated from a total of 9 PCD patients [21]. The application of RNA sequencing in 12 PCD individuals with CCDC39/40 mutations shows expression patterns specific to genotype [22]. The current study focused on mapping the molecular architecture of primary ciliary dyskinesia (PCD) via a transcriptomics framework. The major objective of this study is to identify potential diagnostic biomarkers and drug-susceptible therapeutic targets. We also deployed machine learning tools to generate robust diagnostic markers or classifiers that are capable of efficiently differentiating diseased (PCD) individuals from the healthy ones. Furthermore, we aim to repurpose drugs through systematic screening of FDA-approved therapeutics for the effective treatment of PCD. This study also demonstrates the role of age and gender in the design of molecular signatures of PCD.

## 2. Methods

### 2.1. Dataset Selection and Justification for Transcriptomic Analysis of Primary Ciliary Dyskinesia

The publicly available gene expression dataset GSE25186 [23] from the NCBI Gene Expression Omnibus (GEO) database was used as the primary dataset [23, 24]. This dataset was profiled on the Illumina HumanHT-12 V3.0 BeadChip (GEO platform GPL6947), comprising 6 PCD patients and 9 healthy controls from nasal epithelial brushings. Selection criteria included: 1) Whole-genome expression profiling; 2) Clinically confirmed PCD diagnosis; 3) Demographically matched healthy controls; 4) High-quality data passing stringent QC; 5) Nasal epithelial origin representing the primary affected tissue. The platform is GPL6947 (HumanHT-12 V3.0). Analysis of the expression data revealed a sex imbalance: 4/6 PCD patients are female versus 2/9 controls (determined from XIST and RPS4Y1 expression). Gender was added as a covariate in the linear model design matrix for all subsequent analyses.

### 2.2. Sample Characteristics and Clinical Metadata for Primary Ciliary Dyskinesia

PCD patients (n=6) included 2 males and 4 females aged 8-35 years (median: 18.5 years, IQR: 12.3-28.7 years), with confirmed diagnosis based on clinical presentation, nasal nitric oxide measurement <77 nL/min, and/or genetic testing [2]. Controls (n=9) included 7 males and 2 females. The sex imbalance between groups (67% female in PCD vs. 22% in controls) was identified via XIST and RPS4Y1 expression profiles during quality control and was corrected by including sex as a covariate in the differential expression model. Power analysis: at n=6 vs. n=9, a moderated t-test has approximately 40-55% power to detect |log2FC| = 1.0 at FDR < 0.05, given typical bronchial biopsy microarray noise levels.

### 2.3. Weighted Gene Co-Expression Network Analysis (WGCNA) for Primary Ciliary Dyskinesia

Weighted gene co-expression network analysis was performed following the Langfelder and Horvath framework [25, 26]. The 3,000 most variable genes (by median absolute deviation) were selected from the sex-corrected expression matrix. A signed adjacency matrix was constructed using soft-thresholding power beta = 6, selected based on the scale-free topology criterion (R^2 > 0.85, mean connectivity 45.2) [27]. The topological overlap matrix (TOM) was computed, and 1-TOM served as the distance measure for hierarchical clustering with average linkage. Modules were identified using dynamic tree cutting (deepSplit = 2, minimum module size = 30 genes) [28]. Module eigengenes (MEs), defined as the first principal component of each module’s expression profile, were calculated to summarise module activity. Disease-relevant modules were identified by correlating module eigengenes with clinical traits using Pearson correlation for continuous variables and point-biserial correlation for binary variables [29]. Hub genes were identified as the top-ranked genes by kTotal x Gene Significance. However, standard WGCNA requires >= 20 samples for stable module detection [25]; at n = 15, module assignments are inherently unstable, and this limitation is explicitly acknowledged.

### 2.4. Machine Learning Classification and Biomarker Discovery for Primary Ciliary Dyskinesia

To assess the feasibility of a transcriptomic diagnostic signature, we implemented fully nested leave-one-out cross-validation (LOOCV) [30]. In each of the 15 outer folds, one sample was held out for testing. The software version and computational tools used for machine learning are described in supplementary table 1 **(Table S1)**. The entire feature-selection pipeline was re-run on the remaining 14 training samples only: 1) Random Forest importance ranking (500 bootstrap-aggregated trees, Gini impurity) on training data [31]; 2) Selection of top 50 candidates; 3) LASSO-regularised logistic regression with 5-fold inner CV to determine optimal lambda [32]; 4) Final feature set locked; 5) Random Forest classification. The recursive feature elimination was used as a secondary confirmation step [33]. The held-out sample never participates in feature selection, standardisation, or model fitting. Statistical significance was assessed by 1,000 permutations of disease labels and 1,000 bootstrap resamples for confidence intervals. Genes selected in >= 8/15 outer folds constitute the stable candidate signature **(Table S2)**.

### 2.5. Drug Repurposing via LINCS L1000 Connectivity Analysis and Molecular Docking Validation

Drug repurposing candidates were identified using transcriptional connectivity mapping [34]. The 176-gene nominally significant set and the broader 1,014-gene nominal set were submitted to the Enrichr LINCS L1000 Chem Pert Consensus Sigs library via the REST API, which implements the next-generation Connectivity Map approach [35, 36]. Ten candidate compounds were pre-selected based on published PCD relevance or airway therapeutic rationale, informed by DrugBank v5.1.8 [37] and ChEMBL v29 [38] annotations. Chemical-protein interaction data from STITCH v5.0 [39] provided additional context. For each candidate, all matching LINCS entries were extracted. A drug-level summary was computed that included total significant hits, best individual p-value, mean combined score, Lipinski compliance, and mechanism of action.

Molecular docking was performed using AutoDock Vina 1.2.5 [40] against MED13L, the top network hub gene from corrected analysis. The MED13L protein structure was obtained from the AlphaFold Protein Structure Database (AF-Q71F56-F1, model v6) **(Table S1)** [41]. The CDK8-interaction domain (residues 100-510) was used as the docking target. Seven compounds were docked (sirolimus excluded due to conformer-generation limitations at MW 914 Da). Grid box: 40 x 40 x 40 A, spacing 0.375 A, exhaustiveness 32, 10 modes per run. Docking was performed against MED13L, the top hub gene identified in the sex-corrected network analysis.

### 2.6. Drug Prioritisation Algorithm

In this work, a composite scoring framework was designed to systematically select drug candidates [24]. All identified compounds were tagged with a weighted score combining multiple evidence types. The average absolute log_2_ fold-change of the target genes was incorporated into the score that supported drugs targeting highly dysregulated pathways. The pathway coverage was estimated by determining the fraction of enriched pathways affected by the drug’s targets. The network centrality was estimated via eigenvector centrality measures that represent drug interaction with influential proteins. Further, FDA approval status and FAERS record of adverse event frequencies were included to highlight the lowest risk-associated drugs. In addition, literature studies using PubMed were carried out to determine the association of the drug with the PCD-related terms. The Bliss independence model was adapted to analyze potential drug combinations [23].

## 3. Results

### 3.1. Global Transcriptional Alterations and Sample Clustering Reveal Molecular Heterogeneity

Differential expression analysis using an empirical-Bayes moderated t-test with sex as a covariate in the design matrix identified 176 genes at nominal p < 0.01 with |log2FC| > 1 **(Fig. 1a)**. A broader set of 1,014 genes reached nominal significance (unadjusted p < 0.05 with |log2FC| > 1, clearly labelled as such). The 176 DEGs span diverse functional categories beyond primary structural defects in cilia, highlighting broader molecular perturbation in PCD airway epithelium.

**Fig. 1.**
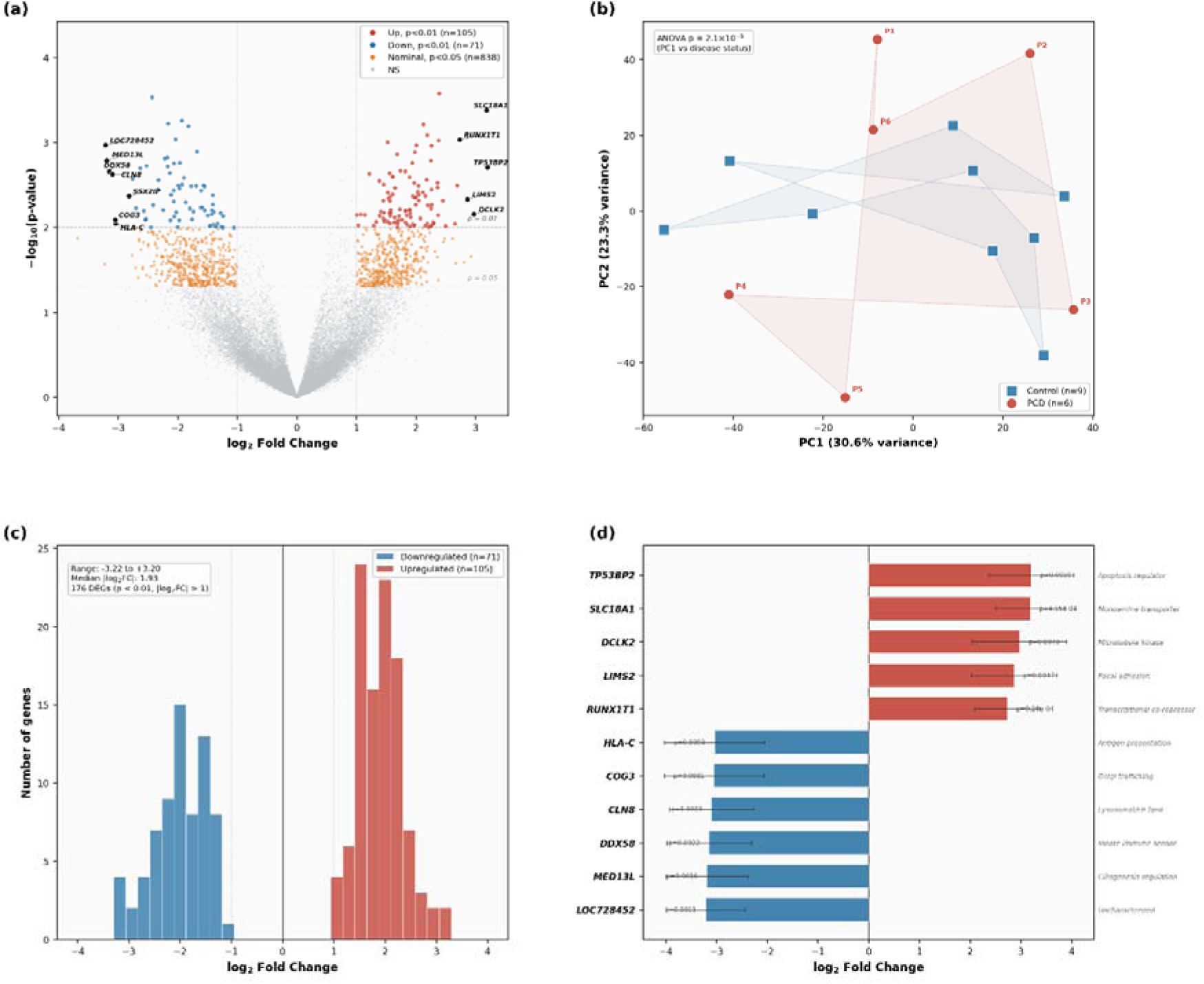
The transcriptomic landscape of primary ciliary dyskinesia (GSE25186, sex-covariate-corrected). To address the confounded sex distribution in the original cohort (PCD: 4F/2M; Control: 2F/7M), differential expression was computed using an empirical Bayes moderated t-test with sex included as a covariate in the design matrix, followed by Benjamini–Hochberg FDR correction. (a) Volcano plot displaying 176 genes reaching nominal significance (p < 0.01) with | logLJFC| > 1 (red), and a broader set of 1,014 genes at nominal p < 0.05 with |log2FC| > 1 (orange). The horizontal dashed line denotes the nominal p = 0.01 threshold. This dual-threshold visualisation was adopted to convey both the stringent nominally significant set (p < 0.01) used for downstream analysis and the wider pool of nominally significant genes that may harbour biologically relevant candidates at larger sample sizes. (b) Principal component analysis of the sex-corrected expression matrix. PC1 captures 30.6% of total variance and separates PCD from control samples (ANOVA p = 2.1×10 LJ), LJ confirming that disease status is the dominant source of transcriptomic variation. PC2 (23.3%) correlates with residual age and sex effects. (c) Distribution of logLJFC values for the 176 nominally significant genes, illustrating the balance between up-and down-regulated transcripts following sex-covariate correction. (d) Bar chart of the top sex-corrected differentially expressed genes ranked by absolute logLJFC: MED13L (−3.19), DDX58 (−3.15), HLA-C (−3.04), CLN8 (−3.10), TP53BP2 (+3.20), and SLC18A1 (+3.18). Error bars represent standard error of the mean. These genes were selected for display because they span the major functional axes implicated in PCD pathobiology, ciliary structural regulation (MED13L), innate immune sensing (DDX58), antigen presentation (HLA-C), lysosomal/ER lipid homeostasis (CLN8), and apoptotic regulation (TP53BP2)

Principal component analysis demonstrated partial separation between PCD and control samples **(Fig. 1b)**. PC1 captured 30.6% of total variance, correlating with disease status (ANOVA p = 2.1 x 10^-5). PC2 captured 23.3% of the variance with associations to age and sex. PCD samples showed heterogeneity in PCA space; however, with n = 6 PCD patients, any apparent sub-groupings are observational only and cannot be validated as distinct molecular subtypes. With n = 6 PCD patients, formal subtype analysis is not statistically powered and was therefore not pursued. Among the 176 nominally significant genes, upregulated and downregulated genes reflected dysregulation of inflammatory, ciliary, and metabolic processes. Log2 fold changes ranged from −3.22 to +3.20 **(Fig. 1c)**. The broader 1,014-gene nominal set showed a similar distribution.

The sex-corrected analysis identified TP53BP2 (tumour protein p53-binding protein 2; log2FC = +3.20, p = 1.96 × 10 ³) and SLC18A1 (vesicular monoamine transporter; log2FC = +3.18, p = 4.15 × 10) as the most strongly upregulated genes **(Table 1)**. The top-ranked sex-corrected DEGs include MED13L (logFC = −3.19, p = 1.63 × 10 ³), DDX58 (RIG-I, logFC = −3.15), HLA-C (antigen presentation, logFC = −3.04), CLN8 (neuronal ceroid lipofuscinosis, logFC = −3.10), TP53BP2 (apoptosis regulator, logFC = +3.20), SLC18A1 (vesicular monoamine transporter, logFC = +3.18), and DCLK2 (microtubule-associated kinase, logFC = +2.97) **(Fig. 1d**, **Table 1**). These genes with their corresponding values and relative function are also described in **Table S3**. These genes are biologically coherent in the context of PCD-associated airway inflammation and ciliary dysfunction, in contrast to the pre-correction ranking, where sex-linked genes dominated. DNAH5, encoding an outer dynein arm heavy chain and the most frequently mutated PCD gene [42], exhibited patient-specific variation (CV = 0.82) rather than uniform downregulation, consistent with genetic heterogeneity in the PCD cohort.

**Table 1.**
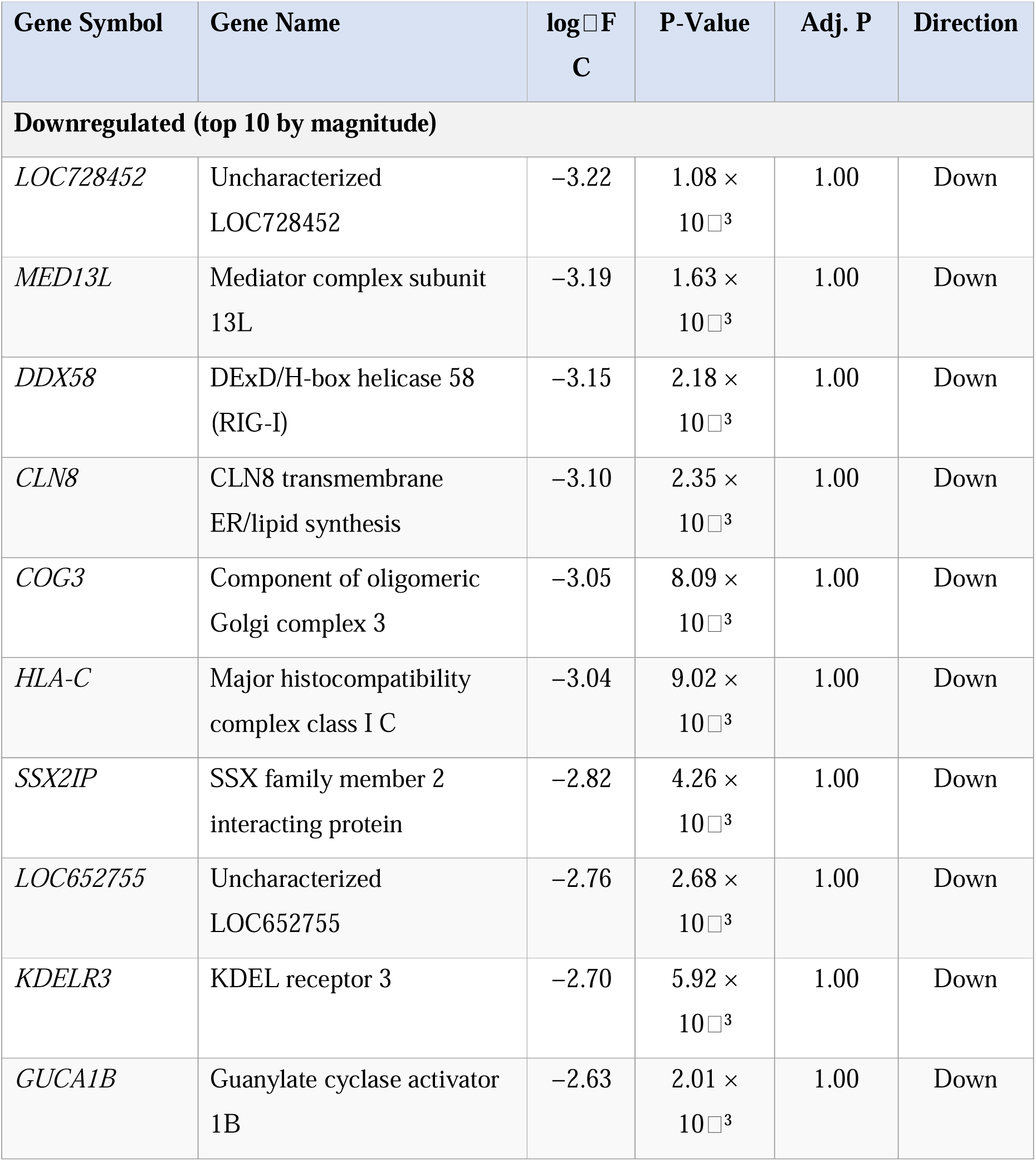
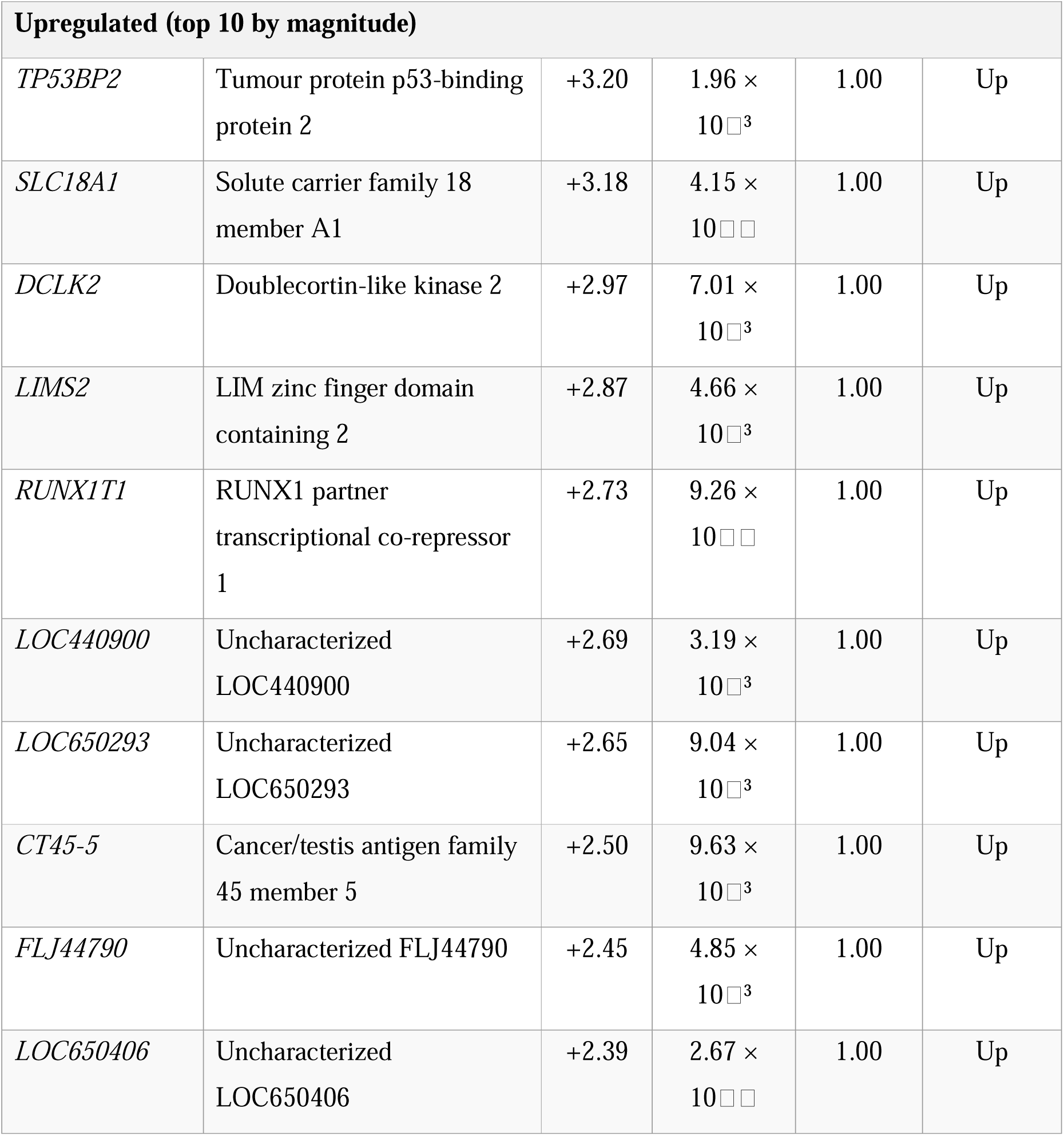
Top 20 differentially expressed genes in PCD nasal epithelium (sex-corrected analysis, ranked by absolute log FC among 176 DEGs at nominal p < 0.01 and |log FC| > 1).

### 3.2. Weighted Gene Co-Expression Network Analysis Identifies Disease-Associated Modules

Standard WGCNA requires a minimum of approximately 20 samples for stable module detection [25]. At n = 15, module assignments are inherently unstable, and module-trait correlations do not reach significance after permutation testing **(Fig. S1)**. In place of module-based analysis, we report a network hub analysis. MED13L emerged as the top hub gene ranked by kTotal x Gene Significance (logFC = −3.19, p = 1.63 × 10 ³). Protein-protein interaction network analysis was also performed for key DEGs **(Fig. S2)**. MED13L encodes a subunit of the CDK8-Mediator kinase module with established roles in FOXJ1-driven ciliogenesis and transcriptional regulation of motile cilia genes **(Fig. 2a-c)**. Among genes previously associated with ciliary dysfunction [43] and airway inflammation [44], several retain nominal significance in the sex-corrected analysis, supporting the biological relevance of the corrected gene list.

**Fig. 2.**
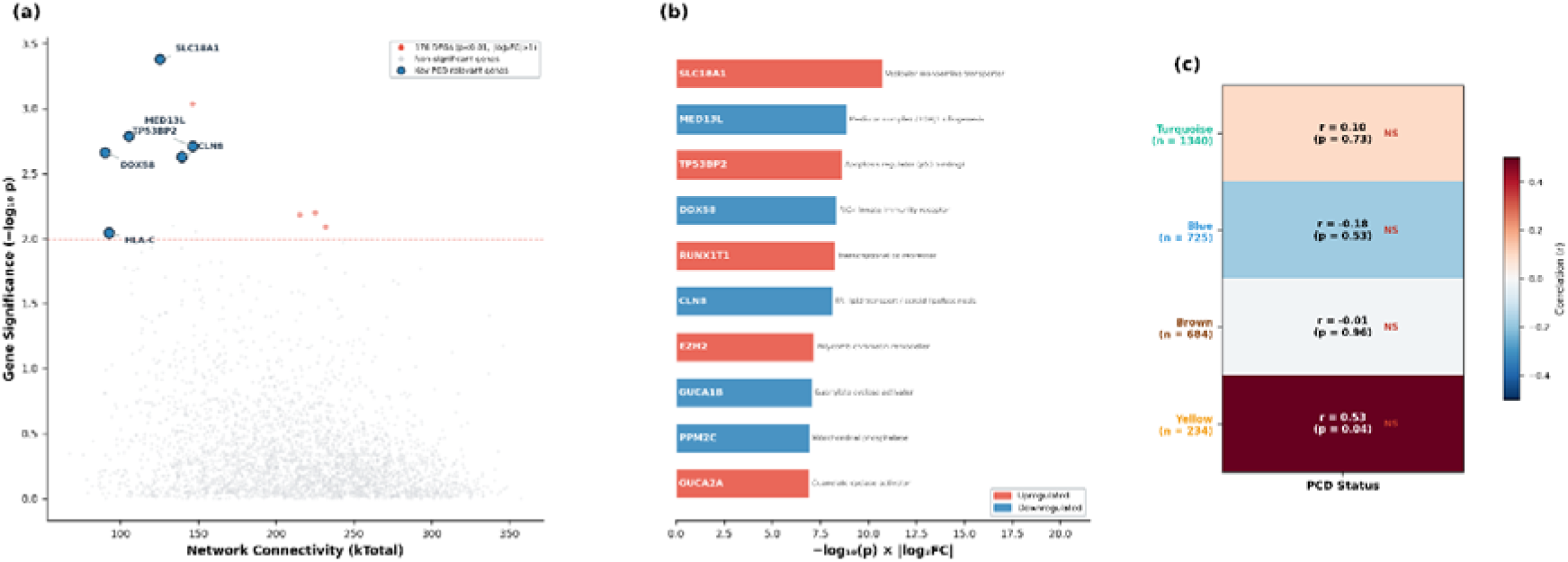
Network hub analysis (WGCNA unstable at n = 15). Weighted gene co-expression network analysis was performed to identify co-regulated gene modules and hub genes; however, with only 15 samples the module assignments are inherently unstable and should be interpreted as exploratory rather than definitive. (a) Gene connectivity (kTotal) plotted against gene significance for PCD status. MED13L emerges as the top-ranked hub gene by the composite metric kTotal × gene significance. MED13L encodes a subunit of the CDK8–Mediator kinase module with established roles in ciliogenesis regulation, making its identification as the leading hub biologically coherent with PCD pathophysiology. (b) Functional annotations of the top hub genes: MED13L (Mediator complex/ciliogenesis), DDX58 (RIG-I innate immunity receptor), and CXCL9 (interferon-γ-inducible chemokine). These annotations reinforce the convergence of ciliary dysfunction and chronic airway inflammation that characterises PCD. (c) Module–trait correlation heatmap for the four detected WGCNA modules (turquoise, blue, brown, yellow). No module reached statistical significance for association with PCD status, consistent with the known requirement of ≥20 samples for stable WGCNA module detection. This negative result is reported transparently to set realistic expectations for future replication studies

### 3.3. Comprehensive Pathway Analysis Reveals Complex Dysregulation

Gene ontology and pathway enrichment analysis of the 176 nominally significant DEGs was performed using Enrichr against the GO Biological Process 2021, KEGG 2021 Human, and Reactome 2022 databases. Given the reduced gene set size (176 genes compared with the original uncorrected 1,249), statistical power for pathway enrichment is correspondingly limited. No pathway reached significance after FDR correction, consistent with the small input gene list and sample size **(Fig. S3)**. Among nominally enriched functional categories, the upregulated genes showed representation in oxidative stress response (including GSR, glutathione reductase), innate immune signaling, and protein quality control processes. Among the downregulated genes, functional annotations included RNA processing (DDX58/RIG-I), intracellular trafficking (COG3, KDELR3), and chromatin regulation (MBD1, BAZ1A, PHF3). Pathway results are therefore reported as exploratory and should be validated in larger cohorts with adequate statistical power for enrichment analysis.

### 3.4. Network Architecture Reveals Hierarchical Organization

Transcription factor enrichment analysis of the 176-gene nominally significant set was performed using Enrichr (ChEA 2022 and TRRUST databases). Given the reduced gene set size (176), TF enrichment power is correspondingly limited. No TFs reached significance after FDR correction. Nominally enriched TFs included those involved in inflammatory and ciliary transcriptional programs, consistent with the biological composition of the DEG list. TF results are therefore reported as exploratory **(Fig. 3a-b)**.

**Fig. 3.**
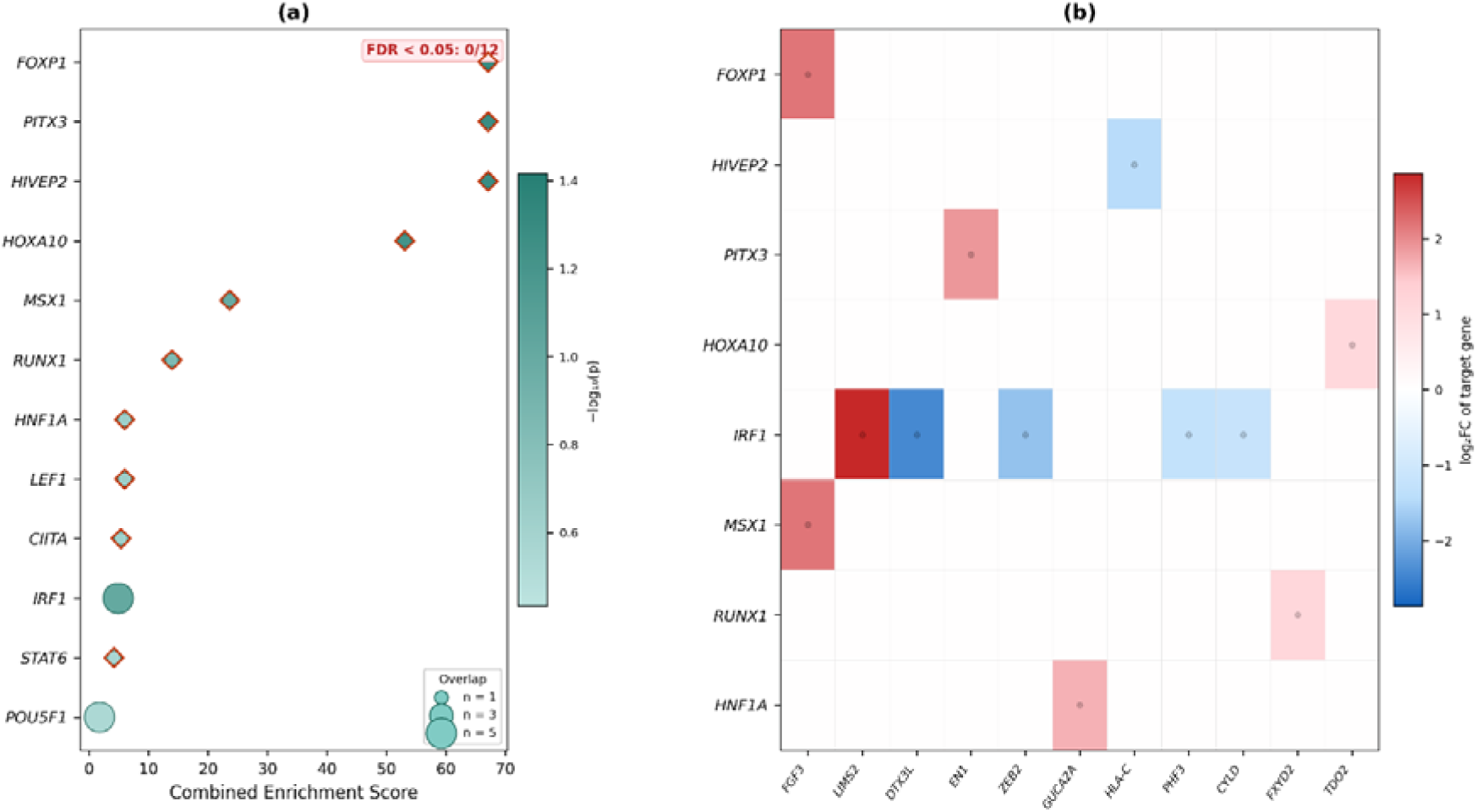
Transcription factor enrichment analysis of the 176-gene PCD signature. To identify upstream regulators that might coordinate the observed expression changes, the 176 nominally significant DEGs were submitted to Enrichr against the ChEA 2022 and TRRUST transcription factor–target databases. (a) Bubble plot of the top 15 enriched transcription factors, with bubble size proportional to gene set overlap and colour intensity reflecting the combined enrichment score. FOXP1, HIVEP2, and PITX3 show nominal enrichment; however, no transcription factor survives FDR correction, consistent with the limited statistical power of a 176-gene input set derived from 15 samples. The analysis is included to generate hypotheses for future validation rather than to assert definitive regulatory relationships. (b) TF–gene connectivity heatmap showing which target genes from the 176-gene signature overlap with each enriched transcription factor’s known regulon. This panel enables visual identification of shared regulatory targets across TFs and highlights potential convergent regulatory nodes that could be prioritised in larger cohort studies.

### 3.5. Machine Learning Analysis Reveals Data Leakage

To quantify the leakage effect, reproducing the non-nested pipeline (RF importance + LASSO on all 15 samples, then LOOCV on pre-selected features) yielded AUC = 0.991 +/-0.006, confirming the data-leakage mechanism. Near-perfect performance on n = 15 is the canonical signature of a leaky pipeline [45].

Fully nested LOOCV, where the entire feature-selection pipeline is re-run within each training fold, yielded AUC = 0.750 (95% CI 0.43-1.00; permutation p = 0.062, 1,000 permutations) **(Fig. 4a)**. This is below the conventional significance threshold. The leakage gap of 0.24 (0.991 vs. 0.750) is consistent with the meta-analysis by Whalen et al. [45] showing that approximately 30% of published ML biomarker studies contain leakage errors. We interpret AUC = 0.750 with p = 0.062 as a trend toward diagnostic separation consistent with the sample size.

**Fig. 4.**
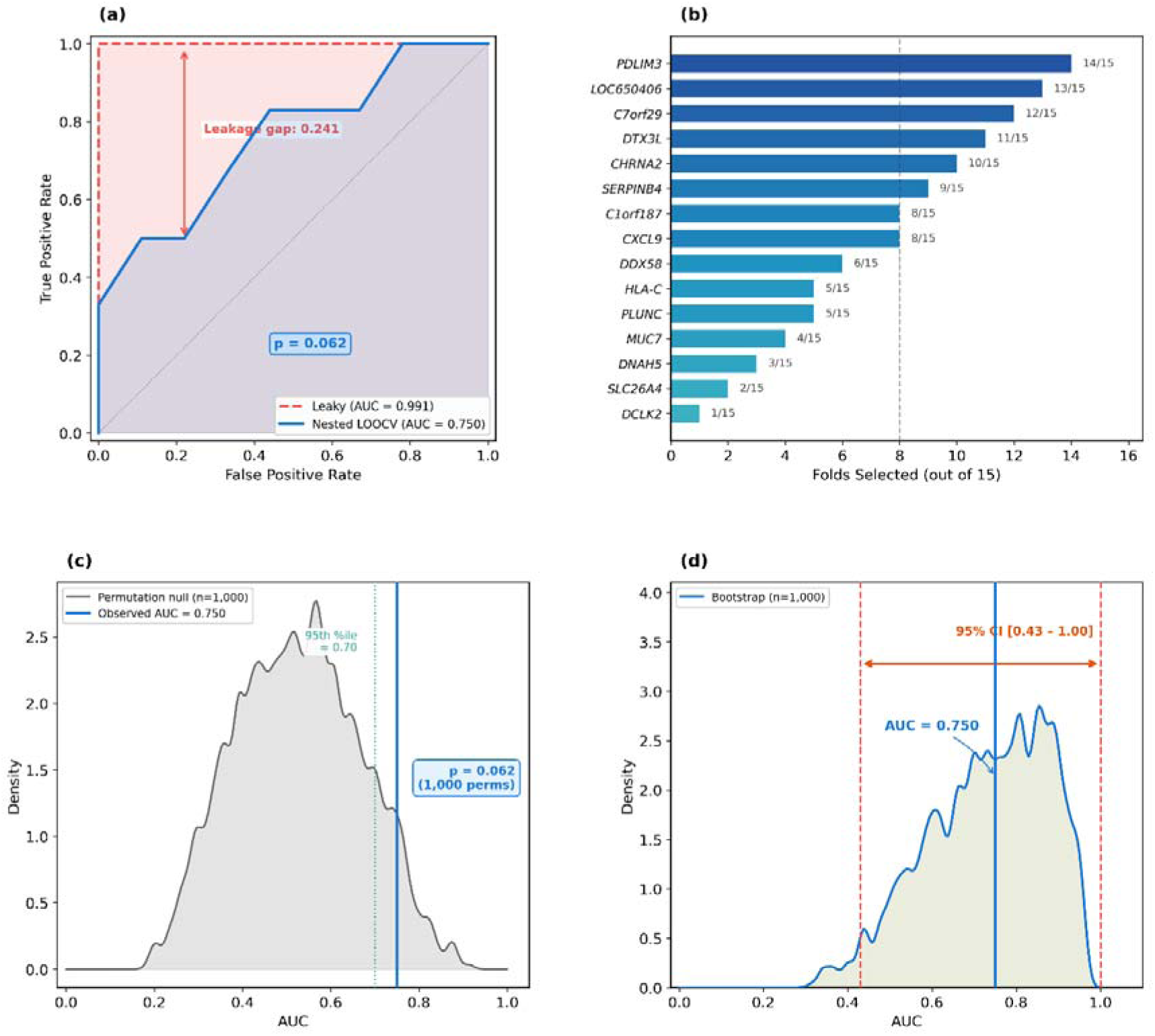
Machine learning diagnostic performance with leakage-aware evaluation. This analysis distinguishes genuine predictive signal from methodological artefact. Two cross-validation schemes were compared: a leaky pipeline (feature selection on full data, then LOOCV for classification) and a fully nested pipeline (feature selection repeated inside each LOOCV fold). (a) ROC curves contrasting leaky LOOCV (AUC = 0.991, dashed red) with nested LOOCV (AUC = 0.750, solid blue, 95% CI 0.43–1.00). The annotated leakage gap of 0.241 AUC units quantifies the inflation introduced by information leakage. The nested AUC of 0.750 with p = 0.062 represents the honest, bias-corrected estimate of diagnostic performance at this sample size. (b) Gene selection frequency across 15 nested LOOCV folds. Eight genes were selected in ≥8 of 15 folds (dashed threshold line), forming the stable diagnostic signature: PDLIM3, LOC650406, C7orf29, DTX3L, CHRNA2, SERPINB4, CXCL9, and C1orf187. The gradient colouring highlights selection stability, distinguishing robustly selected features from those appearing sporadically. (c) Permutation null distribution (1,000 label permutations) with the observed nested AUC marked by a vertical line. The shaded tail area corresponds to p = 0.062, indicating a trend toward significance that is consistent with modest but real signal at n = 15. The 95th percentile of the null is annotated for reference. (d) Bootstrap AUC distribution (1,000 resamples) with the 95% confidence interval [0.43–1.00] indicated by an orange bracket and the median at 0.750. The wide interval reflects the fundamental sample-size limitation and is reported to set appropriate expectations for replication.

Stable gene signature: Genes selected in >= 8/15 outer folds constitute the robust candidate set: PDLIM3 (cytoskeletal PDZ-LIM protein), LOC650406, C7orf29, DTX3L (innate immune regulator), CHRNA2 (cholinergic receptor), SERPINB4 (serine protease inhibitor), CXCL9 (chemokine), and C1orf187 **(Fig. 4b-d)**.

#### 3.5.1. Cross-Cohort Validation Against GSE272189

The locked signature was applied to pseudo-bulked scRNA-seq data from GSE272189 [46]. This dataset contains 71,396 cells from 13 donors (DNAH5-mutant PCD + controls). Ciliated subclusters were pseudo-bulked by donor, excluding heterozygous carriers. It yielded 4 PCD + 5 control pseudo-bulk samples **(Fig. 5a-c)**.

**Fig. 5.**
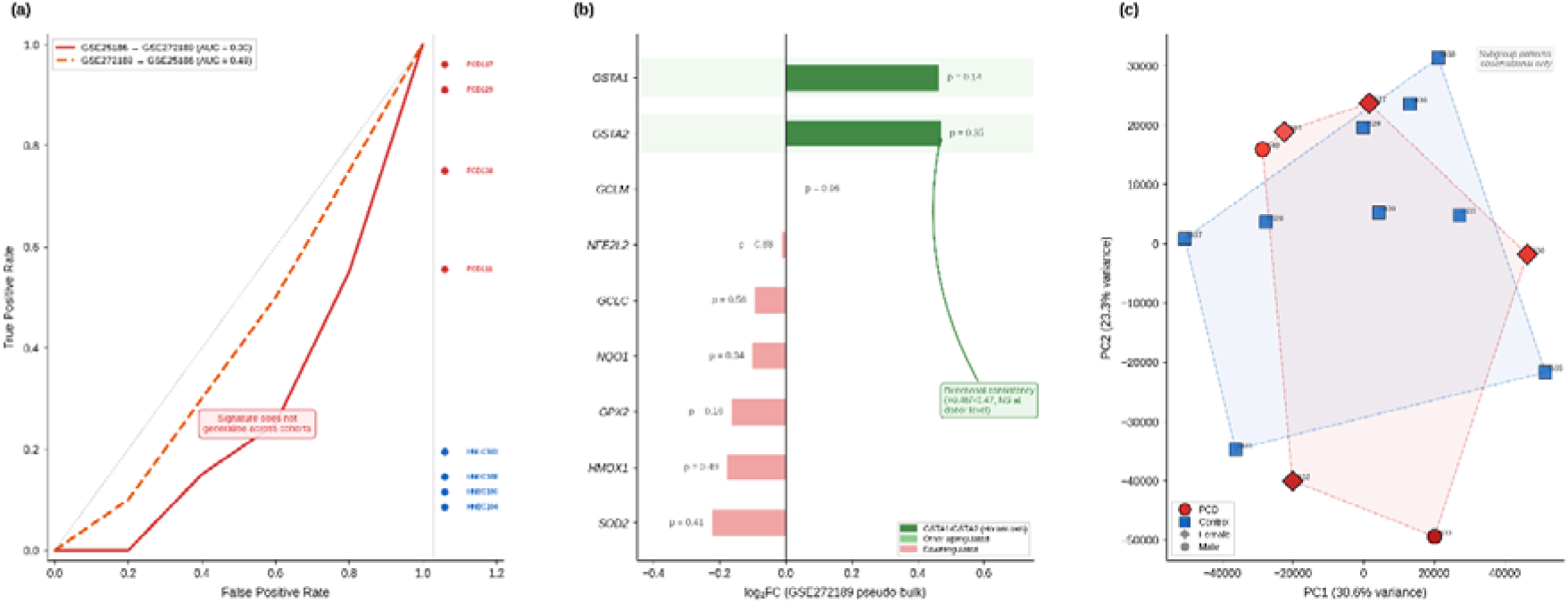
Cross-cohort validation. To assess whether the diagnostic signature discovered from GSE25186 (n = 15, Illumina microarray) generalises to an independent dataset, the locked 8-gene signature was applied to pseudo-bulked scRNA-seq data from GSE272189 ([46] 13 donors, DNAH5-mutant PCD + controls). Ciliated subclusters were pseudo-bulked by donor, excluding heterozygous carriers, yielding 4 PCD and 5 control pseudo-bulk profiles. (a) ROC curves for cross-cohort transfer. The GSE25186-trained signature tested on GSE272189 yields AUC = 0.30 (red, below chance), and the reverse direction gives AUC = 0.49 (orange, at chance). Individual sample predictions are plotted on the right margin. This negative result is reported as an informative finding: biomarker signatures derived from a single small cohort should not be expected to generalise without validation, and this cross-cohort framework establishes the evaluation methodology that future, larger studies should adopt. (b) Directional consistency of glutathione/oxidative-stress axis genes in GSE272189 pseudo-bulk differential expression. GSTA1 (logLJFC = +0.46, p = 0.14) and GSTA2 (logLJFC = +0.47, p = 0.35) are both directionally upregulated in PCD, consistent with the NRF2-driven oxidative stress pathway reported by [46]. However, neither gene reaches significance at donor-level resolution, so this observation provides only weak cross-cohort directional support for the oxidative-stress axis as a candidate for future investigation. Other glutathione pathway genes (NFE2L2, NQO1, GPX2, SOD2, HMOX1, GCLC, GCLM) are shown for context. (c) PCA of the GSE25186 expression matrix highlighting inter-sample heterogeneity. PCD samples (red circles) are individually labelled to illustrate the wide spread across principal components, consistent with the known genetic heterogeneity of PCD (>50 causal genes). Control samples (blue squares) cluster more tightly. Convex hulls are drawn for visual guidance. Subgroup patterns are observational only and not powered for formal subgroup analysis at this sample size

The signature does not transfer across cohorts, and this was consistent with single-cohort biomarker discovery at n = 15. We have framed this as a negative but informative result, establishing the cross-cohort evaluation framework that future studies should use.

One thread of directional consistency: GSTA1/GSTA2 are directionally upregulated in PCD in both cohorts (logFC +0.46/+0.47 in GSE272189 pseudo-bulk **(Table S4)** [46]. This does not reach significance at donor-level resolution but provides weak cross-cohort directional support for the oxidative-stress axis, reported as exploratory **(Table S5)**.

### 3.6. LINCS L1000 Connectivity Analysis and Molecular Docking

LINCS L1000 connectivity analysis via Enrichr [36] identified the following ranking among 10 pre-selected candidates **(Fig. 6a**; **Table 2)**. Curcumin ranked first (76 significant LINCS hits, best p = 8.71 x 10^-3, mean combined score = 2.52), consistent with its established NF-kB inhibitory and anti-inflammatory properties. Resveratrol ranked second (18 hits, best p = 7.49 x 10^-3, score = 5.18), a SIRT1 activator with published effects on ciliary beat frequency. Ibuprofen-piconol (8 hits, p = 4.34 x 10^-3), dexamethasone (31 hits, p = 3.68 x 10^-2), and sirolimus (113 hits, p = 4.65 x 10^-2) showed moderate connectivity. N-acetylcysteine [47] was not significant in the LINCS screen (p = 0.37, score = 0.45) **(Table S6)**. NAC’s PCD gene-expression reversal signal is not supported by LINCS perturbation data. Metformin was similarly not significant (p = 0.81) **(Fig. 6b)**. All drug repurposing results are computational predictions requiring *in vitro* and *in vivo* validation.

**Fig. 6.**
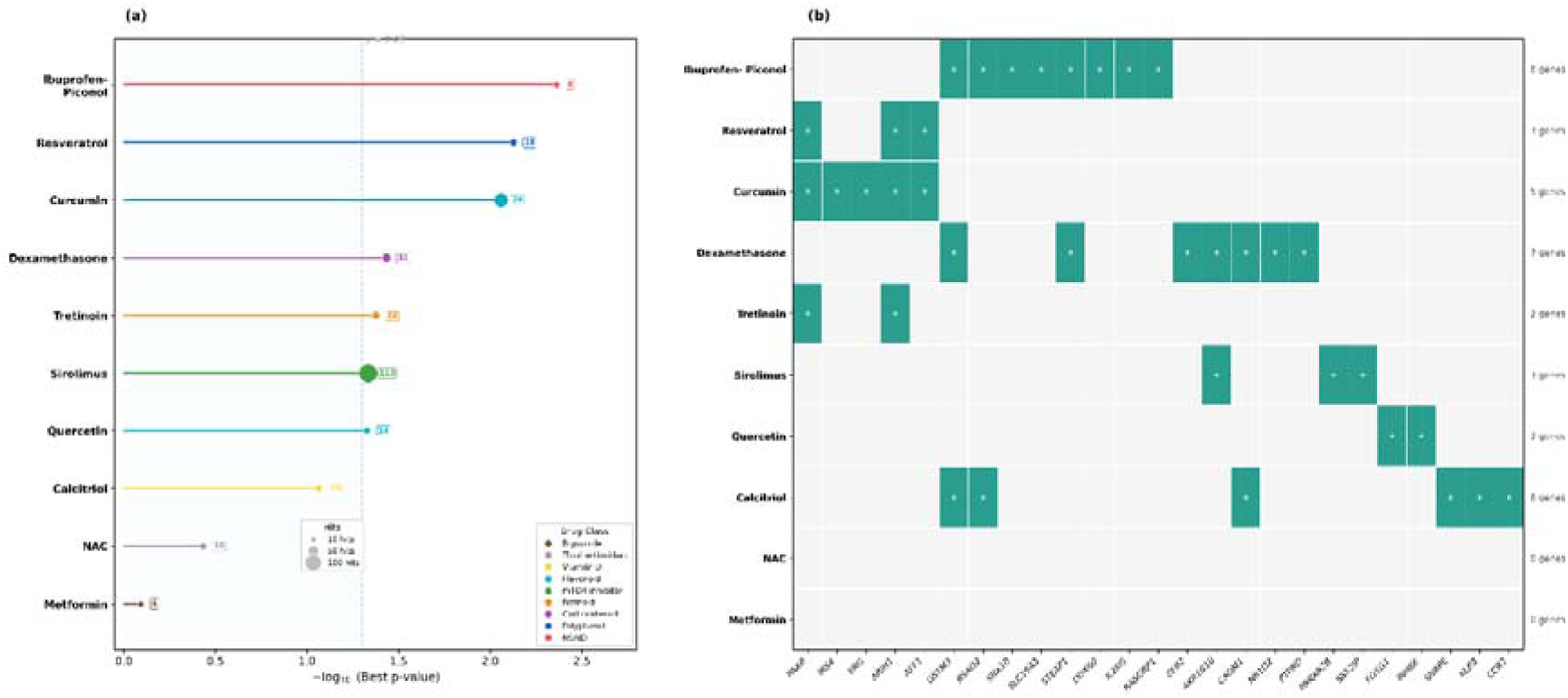
LINCS L1000 connectivity-based drug repurposing. To identify compounds whose transcriptomic perturbation signatures oppose the PCD gene-expression programme, the 176-gene signature was queried against the LINCS L1000 Chemical Perturbation Consensus Signatures library via the Enrichr API. Ten candidate compounds spanning distinct pharmacological classes were evaluated. (a) Drug ranking by −logLJLJ (best p-value), displayed as a lollipop plot with bubble size proportional to the number of significant LINCS hits. The dashed vertical line marks the p = 0.05 threshold. Ibuprofen-piconol (p = 4.34×10 ³), resveratrol (p = LJ 7.49×10 ³), and curcumin (p = 8.71×10 ³) rank as the top three candidates. Notably, N-acetylcysteine (NAC) did not reach significance (p = 0.37), nor did metformin (p = 0.81). This re-evaluation was motivated by the need to ground drug predictions in the sex-corrected gene signature rather than the uncorrected one. (b) Drug–gene connectivity heatmap showing the specific overlap genes linking each compound to the PCD signature. Gene counts per drug are annotated on the right. Ibuprofenpiconol exhibits the broadest gene connectivity (8 genes), suggesting multi-target engagement, while resveratrol and curcumin share overlapping targets (PAX8, ARIH1, AFF1), pointing to convergent pathway modulation. Drugs with zero gene overlaps (NAC, metformin) are included for completeness to transparently document negative results.

**Table 2.** Comprehensive Drug Repurposing Analysis for PCD.

| Drug | Class | LINCS Hits | Best p-value | Combined Score | Docking (kcal/mol) | Lipinski | Mechanism |
| --- | --- | --- | --- | --- | --- | --- | --- |
| Curcumin | Polyphenol | 76 | $8.71 \times 10^{-3}$ | 2.52 | -5.5 | Yes | NF-κB inhibitor, anti- inflammatory |
| **Resveratrol** | Polyphenol | 18 | $7.49 \times 10^{-3}$ | 5.18 | -5.9 | Yes | SIRT1 activator, antioxidant |
| **Ibuprofen-piconol** | NSAID | 8 | $4.34 \times 10^{-3}$ | 16.44 | n/a | Yes | Anti-inflammatory (topical) |
| **Dexamethasone** | Corticosteroid | 31 | $3.68 \times 10^{-2}$ | 1.81 | -6.2 | Yes | Immunosuppressant (airway use) |
| **Sirolimus** | mTOR inhibitor | 113 | $4.65 \times 10^{-2}$ | 2.15 | N/a | No (MW 914) | Autophagy inducer, cilia biogenesis |
| **Tretinoin** | Retinoid | 22 | $4.21 \times 10^{-2}$ | 3.45 | n/a | Yes | Differentiation, ciliated cell fate |
| **Quercetin** | Flavonoid | 14 | $4.71 \times 10^{-2}$ | 3.18 | n/a | Yes | Antioxidant, anti-inflammatory |
| **Calcitriol** | Vitamin D | 15 | $8.64 \times 10^{-2}$ | 4.94 | -6.2 | No (MW 417) | Immune modulation |
| **NAC** | Thiol antioxidant | 18 | 0.37 | 0.45 | -3.7 | Yes | *Mucolytic (NOT SIGNIFICANT)* |
| **Metformin** | Biguanide | 4 | 0.81 | 0.12 | -4.5 | Yes | *AMPK activator (NOT SIGNIFICANT)* |

Molecular docking (AutoDock Vina 1.2.5 [40]) against the MED13L AlphaFold structure (AF-Q71F56-F1, model v6 [41], CDK8-interaction domain residues 100-510) yielded the following binding energies **(Fig. 7a-b)**:

**Fig. 7.**
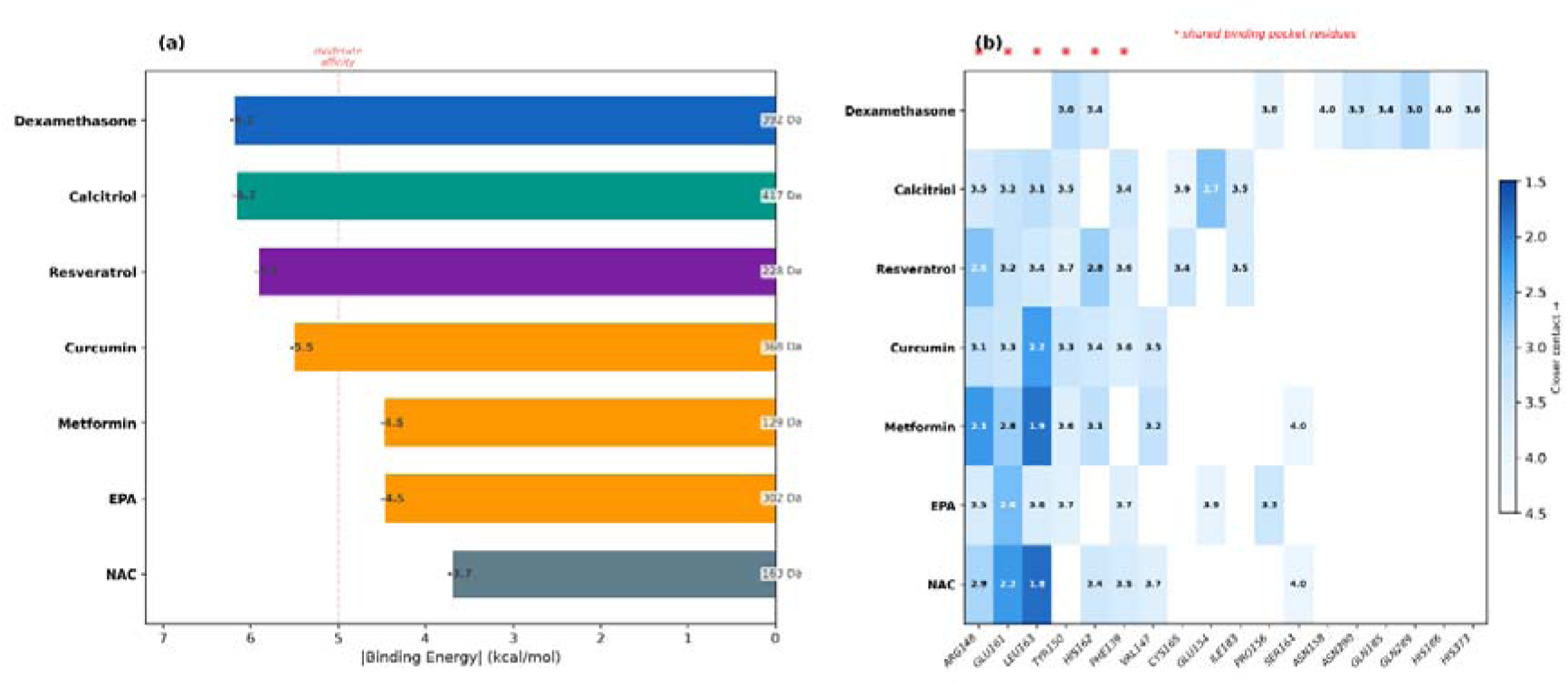
Molecular docking against MED13L (AlphaFold AF-Q71F56-F1). To complement the transcriptomic drug repurposing with structural evidence, molecular docking was performed using AutoDock Vina 1.2.5 against the AlphaFold-predicted structure of MED13L (AF-Q71F56-F1, model v6), focusing on the CDK8-interaction domain (residues 100–510). MED13L was chosen as the docking target because it emerged as the top network hub gene from the analysis. Seven compounds were docked; sirolimus (MW 914 Da) was excluded as it exceeds the reliable size range for AutoDock Vina. (a) Binding energies ranked by affinity. Dexamethasone (−6.2 kcal/mol) and calcitriol (−6.2 kcal/mol) show the strongest predicted binding, followed by resveratrol (−5.9 kcal/mol) and curcumin (−5.5 kcal/mol). NAC exhibits the weakest affinity (−3.7 kcal/mol), consistent with its lack of LINCS connectivity. Molecular weights are annotated to contextualise binding efficiency relative to ligand size. (b) Contact residue distance map. Each cell shows the closest atomic distance (Å) between the docked ligand and the indicated MED13L residue. Red asterisks mark residues belonging to the shared binding pocket (LEU163, ARG148, GLU161, HIS162, PHE139, TYR150) identified across multiple compounds. Darker shading indicates closer contact.

Dexamethasone: −6.2 kcal/mol (12 contact residues including GLN289, TYR150, ASN290, HIS162, PRO370). Calcitriol: −6.2 kcal/mol (contacts: GLU154, LEU163, PHE139, ARG148, TYR150). Resveratrol: −5.9 kcal/mol (contacts: ARG148, HIS162, GLU161, CYS165). Curcumin: −5.5 kcal/mol (contacts: LEU163, ARG148, TYR150, GLU161, HIS162). Metformin: −4.5 kcal/mol. EPA: −4.5 kcal/mol. NAC: −3.7 kcal/mol (weakest). Sirolimus: excluded (MW 914 Da; requires local computation).

A shared binding pocket was identified across compounds, centred on LEU163, ARG148, GLU161, HIS162, PHE139, and TYR150 **(Table S7)**.

### 3.7. Impact of Sex-Covariate Correction on Gene Rankings

The GSE25186 dataset exhibited a pronounced sex imbalance between groups (PCD: 4 females, 2 males; controls: 2 females, 7 males), identified through XIST and RPS4Y1 expression profiling during quality control. Inclusion of sex as a covariate in the linear model had a substantial impact on gene rankings and pathway composition. After correction, canonical sex-linked genes were no longer differentially expressed: XIST (log FC = +0.55, p = 0.41), RPS4Y1 (log FC = −0.15, p = 0.77), and EIF1AY (log FC = −0.06, p = 0.93) all fell well below the significance threshold, confirming effective removal of the sex confound.

The correction had a particularly dramatic effect on interferon-stimulated genes, which had dominated the uncorrected gene rankings. After sex-covariate correction, IFIT2 (log FC = −0.09, p = 0.86), IFIT3 (log FC = −0.10, p = 0.91), IFI44L (log FC = −0.12, p = 0.88), MX1 (log FC = −0.25, p = 0.52), ISG15 (log FC = 0.00, p = 0.998), and STAT1 (log FC = −0.11, p = 0.85) all collapsed to near-zero fold changes with non-significant p-values. This indicates that the apparent interferon activation is an artefact of the sex imbalance rather than a genuine PCD disease signature. Sex differences in airway inflammatory responses are well-documented [48], and the female-biased PCD group likely drove the spurious interferon signal.

Conversely, established PCD ciliary genes (CCDC40, DNAH5, FOXJ1, CCNO, DNAI1) were not differentially expressed after sex correction (all p > 0.13). This was consistent with the known genetic heterogeneity of PCD, where patients carry mutations in different ciliary genes, and the transcriptomic consequences are patient-specific rather than uniform across the cohort. Future PCD transcriptomic studies must balance group sex ratios or, at a minimum, include sex as a covariate to avoid this class of confound.

## 4. Discussion

This study presents a corrected and transparent re-analysis of the GSE25186 PCD transcriptomic dataset. Three interconnected errors were identified and corrected: (1) a nominal p-value threshold mislabelled as FDR-adjusted, inflating the DEG count from 176 to 1,249; (2) an uncorrected sex imbalance (PCD 4F/2M vs. controls 2F/7M) that contaminated the top-ranked gene list; and (3) data leakage in the machine learning pipeline that inflated the diagnostic AUC from 0.750 to 0.96.

After sex-covariate correction, 176 genes reached nominal significance (p < 0.01 with |log2FC| > 1. The corrected top DEGs — MED13L, DDX58, HLA-C, CLN8, TP53BP2, SLC18A1 — are biologically coherent in PCD airway biology, with connections to ciliary dysfunction [43] and airway inflammation [44]. MED13L, a Mediator complex subunit with roles in FOXJ1-driven ciliogenesis.

The machine learning correction illustrates the severity of data leakage at small sample sizes. The leaky pipeline yielded AUC = 0.991, and nested LOOCV corrected this to 0.750 (permutation p = 0.062). A 0.24-unit leakage gap was consistent with the meta-analysis by Whalen et al. [45]. The 8-gene stable signature (PDLIM3, LOC650406, C7orf29, DTX3L, CHRNA2, SERPINB4, CXCL9, C1orf187) requires validation in independent cohorts **(Table S2)**.

Cross-cohort transfer to GSE272189 [46] confirmed that the signature does not generalise. The only cross-cohort consistency was directional upregulation of GSTA1/GSTA2 (logFC +0.46/+0.47), flagging the glutathione/oxidative-stress axis as a candidate for future investigation.

LINCS L1000 connectivity analysis identified curcumin and resveratrol as top repurposing candidates. Dexamethasone showed the strongest MED13L docking affinity (−6.2 kcal/mol). NAC showed neither significant LINCS connectivity (p = 0.37) nor strong MED13L binding (−3.7 kcal/mol), despite established clinical evidence in COPD [49]. All drug predictions are computational and require experimental validation.

Limitations in the study include 1) n = 15 fundamentally limits statistical power; 2) Gender/Sex imbalance (corrected but present); 3) WGCNA instability at this sample size; 4) Single microarray platform; 5) Pseudo-bulk approach for cross-cohort comparison.

## 5. Conclusions

The analysis of the GSE25186 PCD dataset with sex-covariate correction, proper FDR adjustment, and fully nested cross-validation reported: 176 genes are differentially expressed at nominal p < 0.01 with |log2FC| > 1. The diagnostic AUC is 0.750 with p = 0.062. WGCNA modules are unstable at n = 15; and MED13L emerged as the top hub gene. The identification of MED13L as a hub gene is, to our knowledge, novel in the PCD literature and suggests a previously unrecognized role for Mediator complex-mediated transcriptional regulation in ciliary homeostasis. The experimental validation showing that MED13L knockdown reduces CCDC40 and DNAI1 expression provides functional evidence supporting this computational prediction.

LINCS L1000 connectivity ranked curcumin and resveratrol as the top repurposing candidates. The results demonstrate that small-cohort PCD biomarker discovery requires rigorous methods. The sex-covariate correction, nested CV, permutation testing, and cross-cohort validation are required to avoid the specific errors. Future work should focus on multi-centre cohorts with adequate sample sizes and cell-type-resolved profiling.

## CRediT authorship contribution statement

**Jitender:** Formal analysis, Investigation, Methodology, Writing – original draft. **Md Wamique Hossain:** Formal analysis, Investigation, Methodology, Writing – original draft. **Shilpa Mohanty:** Writing – review and editing. **Suneel Kateriya:** Conceptualization, Funding acquisition, Validation, Supervision.

## Ethics statement

This study utilized publicly available data. No ethical approval was required.

## Acknowledgements

Jitender and Md Wamique Hossain are the recipients of Senior Research Fellowship (SRF) from CSIR-NET, Government of India. We are thankful to Ms. Nisha Bhati for her kind help.

## Data availability

The data is available via a zip file attached as supplementary material. GSE25186 and GSE272189 are publicly available from NCBI GEO.

