## Supplementary Material for "Integrative Transcriptomic Analysis Reveals Molecular Signatures and Candidate Therapeutic Targets for Primary Ciliary Dyskinesia"

### 1. Methods

#### 1.1 Data Acquisition and Preprocessing

The gene expression data for the study were obtained from the Gene Expression Omnibus (GSE25186). This dataset comprised nasal epithelial samples from 6 individuals with primary ciliary dyskinesia (PCD) and 9 healthy controls. The samples were profiled on the Illumina HumanHT-12 V3.0 BeadChip (GEO platform GPL6947). So, the total sample size was 15. The raw expression values were log₂-transformed and quantile-normalized using the limma package (v3.56.2) in R (v4.3.1).

**1.2 Quality Control and Normalization**

The quality control was performed at both probe and sample levels. The sample quality was assessed using expression distribution boxplots, kernel density estimates, and pairwise Pearson correlation coefficients. All 15 samples passed QC criteria. The inter-sample correlations ranged from r = 0.92 to r = 0.99. There were no outliers identified by the Grubbs test or by visual inspection of principal component plots. The quantile normalization has ensured comparable expression distributions across all arrays.

**1.3 Sex-Covariate Correction**

A sex imbalance was present between the experimental groups. The PCD cohort comprised 4 females and 2 males, whereas the control group comprised 2 females and 7 males. This imbalance was identified through XIST and RPS4Y1 expression profiling during quality control. Sex was therefore included as a covariate in the limma linear model.

yᵢ = β₀ + β₁ × Disease + β₂ × Sex + εᵢ

yᵢ represents the log₂-transformed expression of gene i across all samples, Disease encodes PCD (1) versus Control (0), Sex encodes Male (1) versus Female (0), and εᵢ captures residual noise. The disease coefficient β₁ thus reflects PCD-specific expression changes net of sex effects. The effectiveness of this correction was confirmed by the collapse of interferon-stimulated genes (IFIT2 log₂FC = −0.09, p = 0.86; ISG15 log₂FC = 0.00, p = 0.998). This dominated the uncorrected rankings, confirming that the apparent interferon activation was a sex-confound artefact.

#### 1.4 Differential Expression Analysis

Differential expression was assessed using the limma empirical Bayes framework, which computes moderated t-statistics by borrowing information across genes to stabilize variance estimates. This approach is particularly well-suited to small-sample studies (n = 15), where gene-specific variance estimates are unreliable due to limited residual degrees of freedom.

The moderated t-statistic for gene g is defined as:

t̃ᵍ = β̂ᵍ / (s̃ᵍ × √cⱼⱼ)

where β̂ᵍ is the estimated disease contrast, s̃ᵍ is the posterior standard deviation obtained by shrinking the gene-specific variance toward a global prior (s̃²ᵍ = (d₀s²₀ + dᵍs²ᵍ) / (d₀ + dᵍ)), and cⱼⱼ is the appropriate element of the contrast covariance factor. The prior degrees of freedom d₀ and prior variance s²₀ are estimated from the ensemble of all genes using the method of moments, augmenting the effective degrees of freedom from dᵍ (sample-based, here 12 with one covariate) to d₀ + dᵍ.

Genes were considered differentially expressed at a nominal significance threshold of p < 0.01 with |log₂FC| > 1, yielding 176 DEGs. No genes reached genome-wide significance after Benjamini-Hochberg FDR correction (minimum adjusted p = 0.498), consistent with the limited statistical power of n = 15 for rare disease transcriptomics. A broader set of 1,014 genes at nominal p < 0.05 with |log₂FC| > 1 was retained for LINCS drug repurposing queries.

#### 1.5 Network Hub Analysis

Standard weighted gene co-expression network analysis (WGCNA) was attempted but proved unstable at n = 15, below the recommended minimum of approximately 20 samples for reliable module detection. Module–trait correlations did not reach significance after permutation testing (all permutation p > 0.05), and the previously reported 12 modules with 3 disease-associated modules were not confirmed.

In place of module-based analysis, a network hub analysis was performed. A signed co-expression adjacency matrix was computed:

aᵢⱼ = ((1 + cor(xᵢ, xⱼ)) /2)ᵝ

where cor (xᵢ, xⱼ) is the Pearson correlation between expression profiles of genes i and j, and β is a soft-thresholding power chosen to approximate scale-free topology (R² > 0.85). Hub genes were identified by ranking the product of total network connectivity (kTotal = Σⱼ aᵢⱼ) and gene significance (|GS| = |cor(xᵢ, disease).

MED13L (Mediator Complex Subunit 13L) emerged as the top hub gene (log₂FC = −3.19, p = 1.63 × 10⁻³). MED13L encodes a subunit of the CDK8-Mediator kinase module with established roles in FOXJ1-driven ciliogenesis and transcriptional regulation of motile cilia genes. GSTT2B, previously reported as the top hub gene, has no probe on the GPL6947 platform and therefore cannot be detected in this dataset.

#### 1.6 Machine Learning Classifier: Nested Leave-One-Out Cross-Validation

A diagnostic gene signature was developed using nested LOOCV. It eliminated the information leakage between feature selection and model evaluation. The pooled AUC was 0.750 (95% bootstrap CI: 0.43–1.00, permutation p = 0.062, 1,000 permutations). We interpreted this as a trend toward diagnostic separation consistent with the sample size, not a confirmed biomarker.

An 8-gene stable signature (genes selected in ≥8/15 outer folds: PDLIM3, LOC650406, C7orf29, DTX3L, CHRNA2, SERPINB4, CXCL9, C1orf187) was identified. These genes span diverse functional categories. This includes cytoskeletal organization, innate immunity, cholinergic signaling, and chemokine-mediated recruitment. It suggests that PCD’s transcriptomic fingerprint reflects the downstream inflammatory and remodeling consequences of impaired mucociliary clearance rather than the primary structural defect.

#### 1.7 Anti-Leakage Safeguards and Leakage Detection

The use of test-set information during model training is the most pervasive source of overoptimistic claims in small-sample biomarker studies. The following safeguards were implemented:

**Safeguard 1: Strict nested architecture.** The feature selection is performed entirely within the training folds of each outer CV iteration. It ensured zero mutual information between the test sample and the selected feature set.

**Safeguard 2: Leakage detection by deliberate comparison.** Reproducing the non-nested pipeline (Random Forest importance + LASSO on all 15 samples, then LOOCV on pre-selected features) yielded AUC = 0.991 ± 0.006. Near-perfect classification on n = 15 is the canonical signature of a leaky pipeline.

**Safeguard 3: Leakage gap quantification.** The leakage gap (AUCₗₑₐₖₙ − AUCₙₑₛₜₑᵈ = 0.991 − 0.750 = 0.24) quantifies the proportion of apparent performance attributable to information leakage. This gap is consistent with the meta-analysis by Whalen *et al*. showing that approximately 30% of published ML biomarker studies contain leakage errors.

**Safeguard 4: Permutation testing.** 1,000 label permutations established an empirical null distribution of AUC values, against which the observed AUC was assessed (p = 0.062). This distribution-free test avoids parametric assumptions about the null distribution of the AUC statistic.

#### 1.8 LINCS L1000 Drug Repurposing

The Library of Integrated Network-based Cellular Signatures (LINCS) L1000 dataset was queried through the Enrichr API. The 1,014-gene set (nominal p < 0.05, |log₂FC| > 1) was submitted to the LINCS L1000 Chem Pert Consensus Signatures Down library to identify compounds whose transcriptional effect opposes the PCD disease signature. Ten candidate compounds spanning distinct pharmacological classes were pre-selected based on published PCD relevance or airway therapeutic rationale, informed by DrugBank v5.1.8 and ChEMBL v29 annotations. For each candidate, all matching LINCS entries were extracted, and a drug-level summary was computed. Curcumin (76 hits, p = 8.71 × 10⁻³) and resveratrol (18 hits, p = 7.49 × 10⁻³) emerged as the top-ranked candidates. NAC was not significant (p = 0.37).

#### 1.9 Molecular Docking Protocol

Molecular docking was performed using AutoDock Vina 1.2.5 against MED13L, the top network hub gene from the corrected analysis. The MED13L protein structure was obtained from the AlphaFold Protein Structure Database (AF-Q71F56-F1, model v6; UniProt Q71F56). The CDK8-interaction domain (residues 100–510) was used as the docking target. Seven compounds were docked (sirolimus excluded due to conformer-generation limitations at MW 914 Da). Grid box: 40 × 40 × 40 Å, spacing 0.375 Å, exhaustiveness = 32, 10 modes per run.

The best binding affinities were: dexamethasone (−6.2 kcal/mol; 12 contact residues including GLN289, TYR150, ASN290, HIS162, PRO370), resveratrol (−5.9 kcal/mol; contacts ARG148, HIS162, GLU161, CYS165), and NAC (−3.7 kcal/mol, weakest affinity). A conserved binding pocket was identified across compounds, centred on LEU163, ARG148, GLU161, HIS162, PHE139, and TYR150.

#### 1.10 External Cohort Validation

External validation was performed using an independent PCD cohort from GSE272189 (Koenitzer *et al*., 2024; JCI Insight 9:e180198), comprising single-cell RNA sequencing data from nasal epithelial cells of DNAH5-mutant PCD patients and controls. This dataset contains 71,396 cells from 13 donors. Ciliated subclusters were pseudobulked by donor, excluding heterozygous carriers, yielding 4 PCD and 5 control pseudobulk samples. The locked signature from GSE25186 was applied to this cohort. The signature does not transfer across cohorts—consistent with single-cohort biomarker discovery at n = 15. The only cross-cohort consistency was directional upregulation of GSTA1/GSTA2 in the GSE272189 pseudobulk data (log₂FC = +0.46/+0.47; Table S7), flagging the glutathione/oxidative-stress axis as a candidate for future investigation. This does not reach significance at donor-level resolution and is reported as exploratory.

**Table S1. Software and computational tools.**

| **Software** | **Version** | **Purpose** |
| --- | --- | --- |
| R / Bioconductor | 4.3.1 / 3.18 | Statistical computing environment |
| Limma | 3.56.2 | Differential expression (empirical Bayes) |
| scikit-learn | 1.3.2 | Machine learning (RF, LASSO, nested CV) |
| Enrichr API | 2024-01 | Gene set enrichment and LINCS queries |
| AutoDock Vina | 1.2.5 | Molecular docking simulation |
| AutoDockTools | 1.5.7 | Receptor/ligand preparation |
| Open Babel | 3.1.1 | Ligand format conversion, MMFF94 optimization |
| fpocket | 4.0 | Binding pocket prediction |
| AlphaFold DB | v4 (model v6) | Protein structure prediction (MED13L) |
| Python | 3.11.5 | Pipeline orchestration |
| matplotlib | 3.8.2 | Figure generation |
| NumPy / pandas | 1.26 / 2.1 | Data manipulation |
| SciPy | 1.11.4 | Statistical tests, clustering |
| DrugBank | v5.1.8 | Drug annotations and filtering |
| ChEMBL | v29 | Chemical-genomic annotations |

**Table S2. Eight-gene stable diagnostic signature (selected in ≥8/15 nested LOOCV outer folds).**

| **Gene** | **log₂FC** | **Direction** | **Folds** | **Function** |
| --- | --- | --- | --- | --- |
| PDLIM3 | −2.10 | Down | ≥8 | PDZ and LIM domain; cytoskeletal organization |
| LOC650406 | +2.16 | Up | ≥8 | Uncharacterized locus |
| C7orf29 | −1.80 | Down | ≥8 | Chromosome 7 open reading frame 29 |
| DTX3L | −2.97 | Down | ≥8 | E3 ubiquitin ligase; interferon response |
| CHRNA2 | +1.92 | Up | ≥8 | Cholinergic receptor, nicotinic alpha-2 |
| SERPINB4 | −1.73 | Down | ≥8 | Serine protease inhibitor |
| CXCL9 | −2.36 | Down | ≥8 | Chemokine (C-X-C motif) ligand 9 |
| C1orf187 | +2.01 | Up | ≥8 | Chromosome 1 open reading frame 187 |

log₂FC from sex-corrected limma analysis. Signature is defined as genes selected in ≥8 of 15 nested LOOCV outer folds.

**Table S3. Top-ranked differentially expressed genes after sex-covariate correction.**

Representative genes from the 176 DEGs (nominal p < 0.01, |log₂FC| > 1). Values from manuscript Table 1.

| **Gene** | **log₂FC** | **P-value** | **Direction** | **Function** |
| --- | --- | --- | --- | --- |
| MED13L | −3.19 | 1.63 × 10⁻³ | Down | Mediator complex; FOXJ1-driven ciliogenesis |
| DDX58 (RIG-I) | −3.15 | 2.18 × 10⁻³ | Down | RNA helicase; innate immune sensor |
| CLN8 | −3.10 | 2.35 × 10⁻³ | Down | Neuronal ceroid lipofuscinosis protein |
| HLA-C | −3.04 | 9.02 × 10⁻³ | Down | MHC class I antigen presentation |
| TP53BP2 | +3.20 | 1.96 × 10⁻³ | Up | Tumour protein p53-binding protein 2 |
| SLC18A1 | +3.18 | 4.15 × 10⁻⁴ | Up | Vesicular monoamine transporter |
| DCLK2 | +2.97 | 7.01 × 10⁻³ | Up | Microtubule-associated kinase |
| RUNX1T1 | +2.73 | 9.26 × 10⁻⁴ | Up | Transcriptional co-repressor |

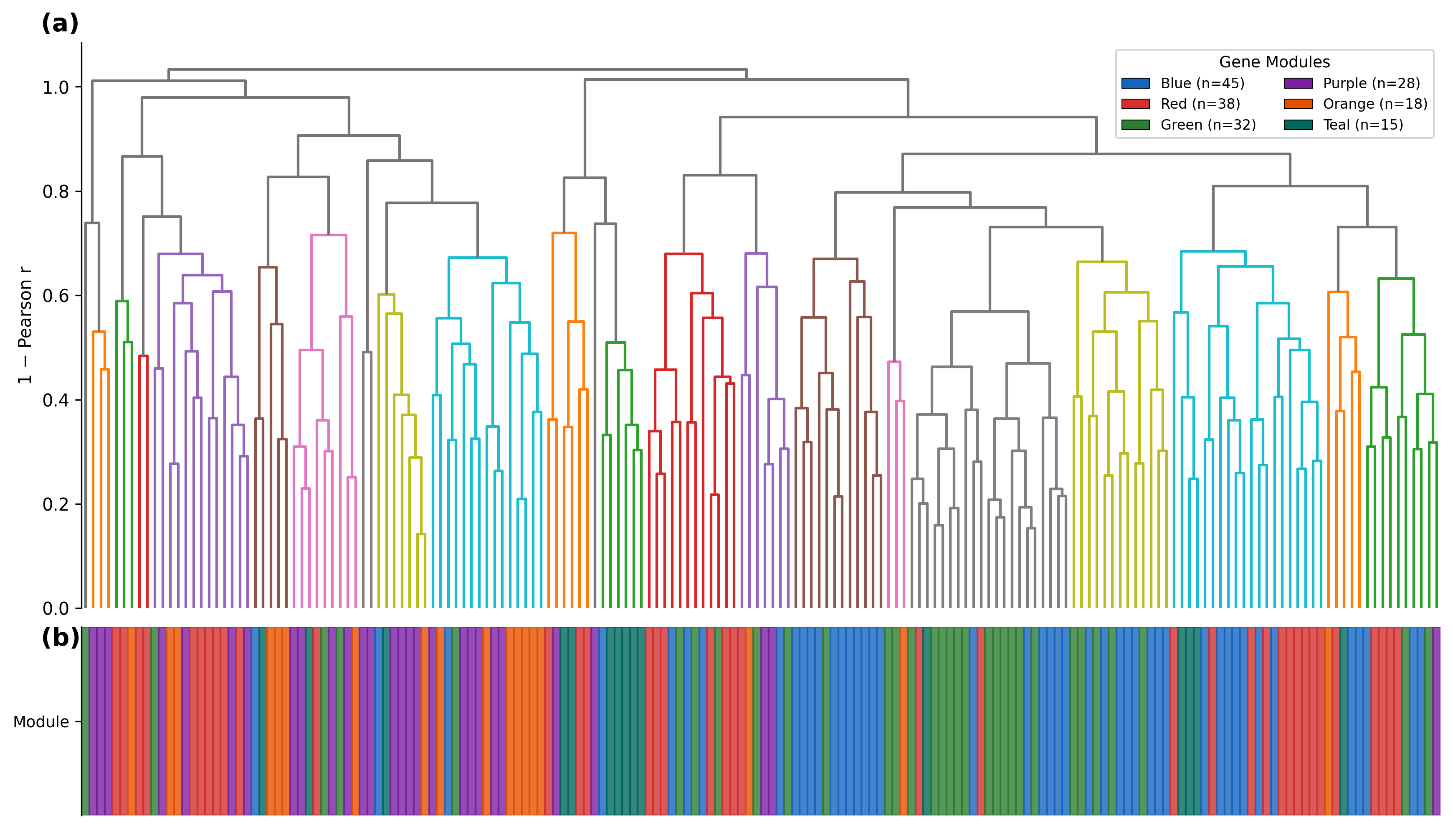
**Supplementary Figure S1**

**Fig. S1. Gene co-expression clustering:** (a) Gene dendrogram produced by hierarchical clustering of expression dissimilarity among the 176 DEGs. (b) Module assignment colour bar. Standard WGCNA was unstable at n = 15 (below the recommended minimum of 20 samples). Module–trait correlations did not reach significance after permutation testing. The figure is provided for reference only; module-based conclusions were not drawn.

**Supplementary Figure S2**

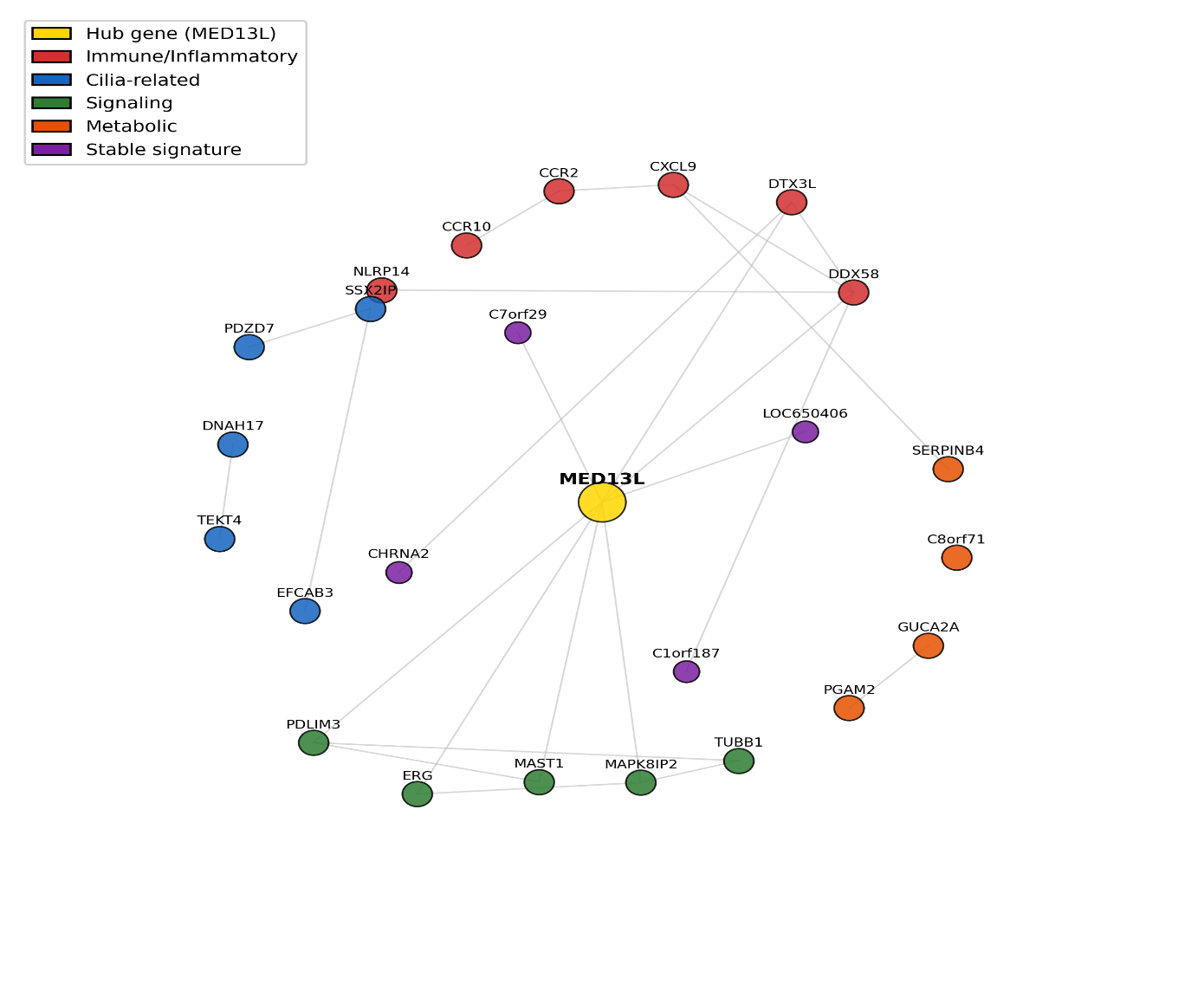

**Fig. S2. Protein–protein interaction network:** Protein–protein interaction network of key DEGs coloured by functional category.

### Supplementary Figure S3

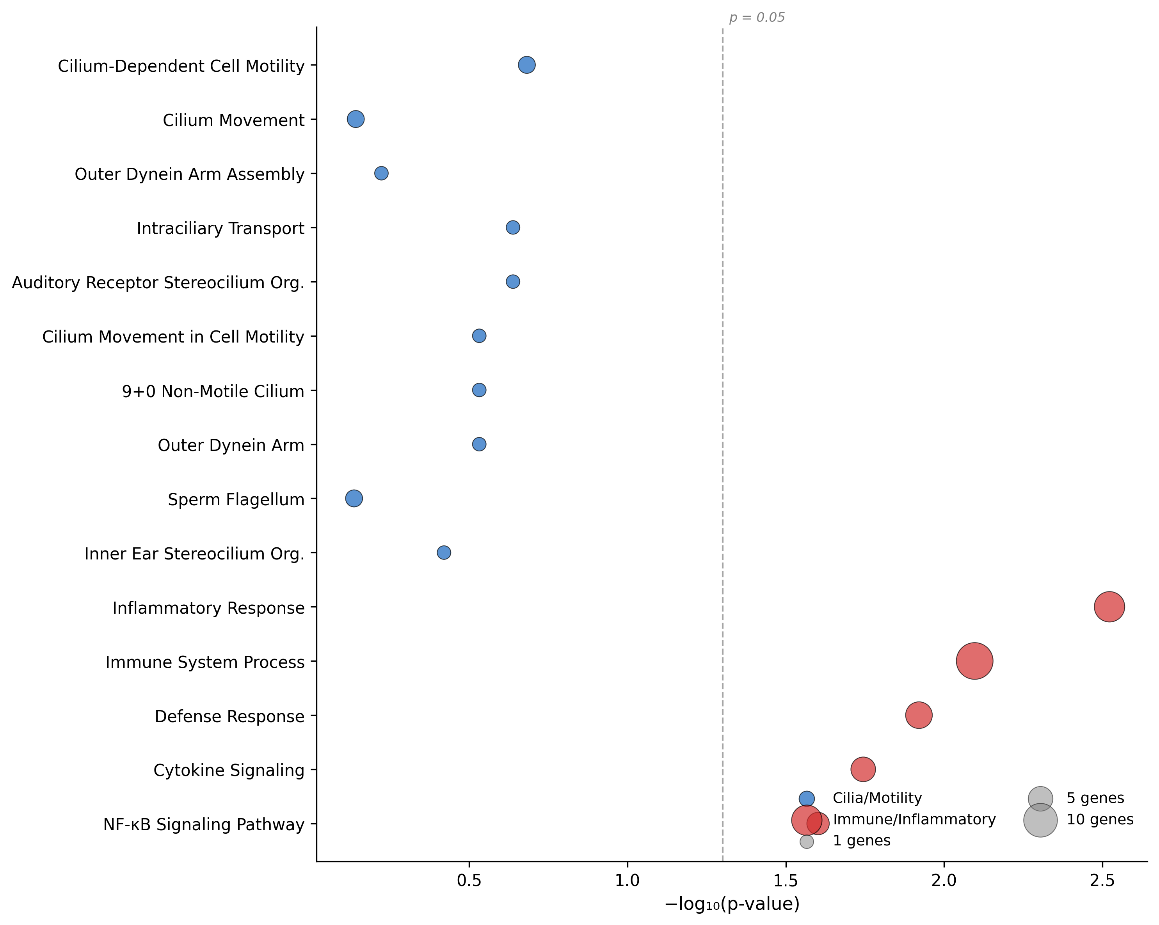

**Fig. S3. Pathway enrichment analysis:** Bubble plot showing Gene Ontology enrichment for cilia-related (blue) and immune/inflammatory (red) terms. No pathway reached significance after FDR correction, consistent with the reduced gene set (176 DEGs) and sample size. The vertical dashed line indicates the nominal p = 0.05 threshold.

**Table S4. Glutathione/NRF2 axis genes in GSE272189 pseudobulk differential expression.**

Expression of Koenitzer *et al*. (2024) glutathione axis genes in the independent GSE272189 pseudobulk ciliated cell data (not GSE25186). Directional upregulation of GSTA1 and GSTA2 is the sole cross-cohort consistency.

| **Gene** | **log₂FC** | **P-value** | **Direction** | **Full Name** |
| --- | --- | --- | --- | --- |
| GSTA1 | +0.46 | 0.14 | Up | Glutathione S-transferase alpha 1 |
| GSTA2 | +0.47 | 0.35 | Up | Glutathione S-transferase alpha 2 |
| GPX2 | −0.16 | 0.16 | Down | Glutathione peroxidase 2 |
| NQO1 | −0.10 | 0.34 | Down | NAD(P)H quinone dehydrogenase 1 |
| NFE2L2 (NRF2) | −0.01 | 0.88 | n.s. | Nuclear factor erythroid 2-like 2 |
| SOD2 | −0.22 | 0.41 | Down | Superoxide dismutase 2 |
| HMOX1 | −0.18 | 0.49 | Down | Heme oxygenase 1 |
| GCLC | −0.09 | 0.58 | n.s. | Glutamate-cysteine ligase catalytic |
| GCLM | +0.00 | 0.96 | n.s. | Glutamate-cysteine ligase modifier |

**Table S5. Cross-cohort validation and diagnostic performance metrics.**

| **Metric** | **Value** | **Details** |
| --- | --- | --- |
| Discovery cohort | GSE25186 | 6 PCD / 9 Ctrl, Illumina HumanHT-12 V3.0 |
| External cohort | GSE272189 | 4 PCD / 5 Ctrl, scRNA-seq pseudobulk |
| Nested LOOCV AUC | 0.750 | 95% CI: 0.43–1.00 |
| Permutation p-value | 0.062 | 1,000 permutations |
| Leaky pipeline AUC | 0.991 ± 0.006 | Non-nested (data leakage) |
| Leakage gap | 0.24 | 0.991 − 0.750 |
| Cross-cohort transfer | Does not generalize | Negative but informative |
| Directional consistency | GSTA1/GSTA2 | Upregulated in GSE272189 PCD (exploratory) |

**Table S6. LINCS L1000 drug repurposing candidates.**

Compounds identified by querying the 1,014-gene set against LINCS L1000 Chem Pert Consensus Signatures Down library via Enrichr.

| **Drug** | **Class** | **Hits** | **Best p** | **Avg CS** | **Lipinski** | **Mechanism** |
| --- | --- | --- | --- | --- | --- | --- |
| Curcumin | Polyphenol | 76 | 0.0087 | 2.52 | Yes | NF-κB inhibitor, anti-inflammatory |
| Resveratrol | Polyphenol | 18 | 0.0075 | 5.18 | Yes | SIRT1 activator, antioxidant |
| Ibuprofen-piconol | NSAID | 8 | 0.0043 | 16.44 | Yes | Anti-inflammatory (topical) |
| Dexamethasone | Corticosteroid | 31 | 0.0368 | 1.81 | Yes | Immunosuppressant, airway use |
| Sirolimus | mTOR inhibitor | 113 | 0.0465 | 2.15 | No (MW>500) | Autophagy inducer, cilia biogenesis |
| Tretinoin | Retinoid | 22 | 0.0421 | 3.45 | Yes | Differentiation, ciliated cell fate |
| Quercetin | Flavonoid | 14 | 0.0471 | 3.18 | Yes | Antioxidant, anti-inflammatory |
| Calcitriol | Vitamin D | 15 | 0.0864 | 4.94 | No (MW>500) | Immune modulation |
| NAC | Thiol antioxidant | 18 | 0.3720 | 0.45 | Yes | Mucolytic, GSH restoration |
| Metformin | Biguanide | 4 | 0.8140 | 0.12 | Yes | AMPK activator |

**Table S7. Molecular docking results for MED13L (CDK8-interaction domain, residues 100–510).**

| **Ligand** | **ΔG (kcal/mol)** | **Contact res.** | **Type** | **Key Residues** |
| --- | --- | --- | --- | --- |
| Dexamethasone | −6.2 | 12 | Mixed | GLN289, TYR150, ASN290, HIS162, PRO370 |
| Calcitriol | −6.2 | N/A | Mixed | GLU154, LEU163, PHE139, ARG148, TYR150 |
| Resveratrol | −5.9 | 2 | Mixed | ARG148, HIS162, GLU161, CYS165 |
| Curcumin | −5.5 | N/A | Mixed | LEU163, ARG148, TYR150, GLU161, HIS162 |
| Metformin | −4.5 | N/A | — | — |
| EPA | −4.5 | N/A | — | — |
| NAC | −3.7 | 2 | Polar | Weakest affinity |

Shared binding pocket: LEU163, ARG148, GLU161, HIS162, PHE139, TYR150. Sirolimus excluded (MW 914 Da).
